# Analysis of potential iron toxicity in hemodialysis patients under intravenous iron treatment

**DOI:** 10.1101/2023.12.05.23299516

**Authors:** Jessy Korina Peña-Esparragoza, Alina Chávez Guillén, Paloma Ramos López, Oscar Rueda Elías, Susana López Ongil, Matilde Alique, Rafael Ramírez-Chamond, Julia Carracedo, Diego Rodríguez-Puyol, Patricia Martínez Miguel

## Abstract

**Background:** The use of higher doses of iron for the treatment of anemia in hemodialysis patients allows lower doses of erythropoiesis-stimulating agents; however, there are concerns regarding the risk of iron toxicity. This study aimed to evaluate the potential toxicity of iron deposition in prevalent hemodialysis patients on iron therapy and its relationship with parameters used to assess iron status, plasma protein oxidation, and cellular iron toxicity.

**Methods:** Magnetic resonance imaging was performed in 56 patients to assess hepatic iron deposition, which was related to clinical and analytical parameters. In patients included in the first and fourth quartiles according to hepatic iron deposition, plasma protein oxidative stress was quantified, as well as iron and cytokine levels in peripheral blood mononuclear cells (PBMCs).

**Results:** Patients with higher hepatic iron deposition had a longer time on hemodialysis (41.0±44.9 vs 4.9±3.4 months, p<0.001) and higher ferritin levels (1181±532 vs 429±278 ng/ml, p<0.001) than those with lower hepatic iron deposition, without differences in transferrin saturation or hepatic enzyme serum concentration. No differences were found in plasma protein oxidation, iron content, or cytokine mRNA content in PBMCs, except for a decrease in IL-6 levels in patients with higher hepatic iron deposition.

**Conclusions:** Patients with longer hemodialysis times had higher iron stores, suggesting that iron treatment over time increases hepatic iron deposition. No parameters supporting increased toxicity in patients with higher hepatic iron deposition were observed; therefore, more proactive treatment with intravenous iron to improve anemia management may not necessarily induce deleterious effects in hemodialysis patients.

## Introduction

In patients receiving maintenance hemodialysis, intravenous iron has become a standard treatment for the management of anemia ^(1)^. Iron is an essential element in several metabolic routes, with particular significance as a heme group compound, and iron supplementation is regularly used even without severe iron deficiency in order to decrease exposure to erythropoiesis-stimulating agents ^(2–3)^.

The parameters currently available to assess iron deficiency in hemodialysis patients are mainly serum transferrin saturation (TSAT) and serum ferritin. Iron deficiency is considered to be either absolute (TSAT<20% and ferritin<100 µ/l) or functional (TSAT<20% and ferritin>200 µ/l). In the latter, iron is sequestered and unavailable for erythropoiesis. However, experts believe that these parameters are not completely reliable for estimating body iron stores or predicting response to therapy, and this is an area that requires additional research ^(4–6)^.. Additionally, it is well known that the improvement of anemia is more effective as iron dose increases; however, there are concerns regarding its potential toxicity. ^(7)^.

Iron is an essential element for the development of infectious microorganisms, and it is believed that excess iron may increase the risk of infections ^(8–9)^..In addition, iron is involved in oxidation-reduction reactions, which can lead to oxidative stress. This may be a contributing factor to increased cardiovascular risk and damage to tissues and organs ^(10–11)^..In fact, retrospective observational studies suggest an increased risk of infections and mortality with high-dose iron administration ^(12)^.This is believed to be particularly relevant when iron stores are high; therefore, the KDIGO Anemia 2012 guideline proposes that iron treatment be avoided when ferritin is above 500 µg/l. However, a recent prospective clinical trial has shown that there is no difference in the incidence of mortality, non-fatal cardiovascular events, and infections between patients treated with proactive(264 mg/monthly) or reactive (145 mg/monthly)doses of iron, considering a limit for ferritin levels higher than those recommended by the KDIGO guidelines (700 μg/l). In this previous study, patients treated with higher iron doses required fewer doses of erythropoiesis-stimulating agents and fewer blood transfusions ^(13)^. Additionally, another recent meta-analysis failed to demonstrate a higher risk of infections or cardiovascular events in patients treated with intravenous iron during dialysis^(14)^. Therefore, there is controversy regarding the maximum dose of intravenous iron that can be used in hemodialysis patients and the levels of ferritin that can be assumed to avoid toxicity.

For this purpose, we aimed to evaluate the potential toxicity of iron deposition in prevalent hemodialysis patients on intravenous iron therapy and its relationship with the parameters used to assess iron status (ferritin and TSAT). In this study, we selected liver and peripheral blood mononuclear cells (PBMCs) to simultaneously test iron deposition and cell toxicity. We also evaluated plasma protein oxidation as a marker of increased oxidative stress.

## Materials and methods

### Study design and population

This cross-sectional study included 56 patients with end-stage renal disease. Patients were aged over 18 years, undergoing in-center thrice-weekly hemodialysis for at least six months, and treated with intravenous iron sucrose. Patients with documented liver disease or cancer were excluded. Moreover, the prevalence of hypertension, diabetes, primary cause of kidney failure, and vascular access were registered. This study was conducted in accordance with the principles of the Declaration of Helsinki and was approved by the Research Ethics Committee (CEIm) of the University Hospital Príncipe de Asturias (HUPA) from Alcalá de Henares (Madrid, Spain).

The data for this study was collected in March 2020 and an informed consent was obtained from all patients included.

Magnetic resonance imaging (MRI) was performed in all patients. In the same month as the MRI, pre-dialysis blood samples were collected in the middle of the week for the analyses detailed below. The mean weekly doses of intravenous iron sucrose and erythropoiesis-stimulating agents in the last 3 months were quantified.

### Magnetic resonance imaging

The MRI signal intensity was measured in five regions of approximately 1 cm^2^. Three were measured in the liver parenchyma, excluding vascular structures, and one was measured in each paraspinal muscle. This process was repeated for the five MRI sequences, and these^(^ values were used to compute five different liver-to-muscle signal intensity ratios, which were then analyzed using the algorithm developed by Gandon et al. ^(15)^ and provided on the website of the University of Rennes ^(16)^.

### Laboratory analysis

#### Evaluation of liver function, anemia, iron metabolism-related parameters, and inflammatory status

Routine blood tests, including measurements of hemoglobin, serum ferritin, transferrin, iron, transaminase, alkaline phosphatase, and C-reactive protein, were performed. Serum levels of alkaline phosphate, aspartate aminotransferase, alanine aminotransferase, gamma-glutamyl transferase, iron, and C-reactive protein were determined by a colorimetric method using an Atellica® analyzer (Siemens Healthineers, Tarrytown, NY, USA), while blood hemoglobin levels were determined by a dual method using a hematology autoanalyzer (Advia 2120i Hematology Systems; Siemens Healthineers, Erlangen, Germany). Moreover, ferritin was measured by chemiluminescence and transferrin was measured by immunoturbidimetry using an Atellica® Analyzer (Siemens Healthineers, Tarrytown,NY, USA).

#### Quantification of iron content in peripheral blood mononuclear cells

An iron assay kit (ab83366, ABCAM, Cambridge, MA, USA) was used to quantify the total Fe, Fe^2+^, and Fe^3+^in the PBMCs. Cells (10^3^/ml) were homogenized in an iron assay buffer on ice and subsequently centrifuged at 16,000 x g to remove insoluble materials. The supernatants were then transferred to a clean tube and kept on ice. Thereafter, 25 µL of the sample was added to a 75 µL iron assay buffer and 5 µL iron reducer, which reduced Fe^3+^ to Fe^2+^. Next, 100 µL of the iron-probe solution was added to generate an Fe^2+^-ferrene S complex that absorbs light at 593 nm, and spectrophotometry was used to detect the absorbance at this wavelength. To normalize the results, the protein content of the samples was calculated and expressed as nmol Fe/mg protein.

#### Evaluation of proinflammatory cytokine mRNA content in PBMCs

Total RNA was extracted from PBMCs using the QIAcube standard protocol (RNeasy QIAcube Kit; QIAGEN) according to the manufacturer’s instructions. Moreover, cDNA was synthesized using the High-Capacity cDNA Archive Kit (Applied Biosystems, Foster City, California, USA)and 1 µg of total RNA primed with random hexamer primers, using a T100 PCR thermal cycler (Bio-Rad, Hercules, California, USA), according to the manufacturer’s instructions. Real-time polymerase chain reaction (PCR) was performed using the ABI Prism 7,500 sequence detection PCR system (TaqMan® Universal Master Mix II, No AmpErase® UNG; Applied Biosystems), according to the manufacturer’s protocol. Additionally, specific TaqMan assay probes from humans used CCL2 or monocyte chemoattractant protein-1 (MCP1) (Hs00234140_m1), tumor necrosis factor alpha (TNF-α) (Hs00174128_m1), interleukin 6 (IL- 6)(Hs00174131_m1), and HPRT1(Hs02800695_m1; normalized assay). The mRNA copy numbers were calculated for each sample using the instrument software and a comparative threshold (Ct) value. After normalization to internal controls (HPRT1 expression levels), the relative fold-change was determined using the 2−ΔΔCt method. Furthermore, gene fold expression was calculated from the difference between the gene expression under each experimental condition.

#### Evaluation of hydroxynonenal protein adducts in plasma

Hydroxynonenal (HNE) protein adducts, an accepted marker of oxidative stress ^(17)^, were measured in the plasma using an OxiSelect HNE adduct competitive ELISA Kit (STA-838) from Cell Biolabs, Inc. (San Diego, CA, USA). This ELISA kit is an enzyme immunoassay developed for the rapid detection and quantitation of HNE protein adducts. The ELISA plate was coated with HNE conjugate, and unknown HNE protein samples from plasma or HNE-BSA standards were added to the HNE-conjugate preabsorbed ELISA plate. After a brief incubation period of 10 min at R/T, an anti-HNE polyclonal antibody was added for 1h at R/T, followed by an HRP-conjugated secondary antibody for 1h at R/T. The content of HNE protein adducts in unknown samples was determined by comparison with a predetermined HNE-BSA standard curve after reading the absorbance at a wavelength of 450 nm, following the manufacturer’s instructions.

### Statistical analyses

Continuous variables are presented as mean ± standard deviation or median (interquartile range), and categorical variables are presented as frequencies and percentages. Continuous and categorical variables were compared using ANOVA and the Chi-square test, respectively. All analyses were conducted using the statistical software IBM® SPSS® Statistics 20, and statistical significance was defined as a two-sided p-value<0.05.

## Results

Hepatic iron deposition was measured by MRI in 56 patients. These patients were divided into four categories, from normal to severe hepatic iron deposition, according to the Rennes score. The baseline patient characteristics are shown in Table 1. Most patients presented with slight iron deposition (41– 100 mOsm/g), and patients with higher hepatic iron deposition had a longer time on hemodialysis and higher serum ferritin levels. No statistically significant differences were observed in the other parameters that were compared, including liver enzyme activity or markers of systemic inflammation such as C-reactive protein.

**Table 1.**
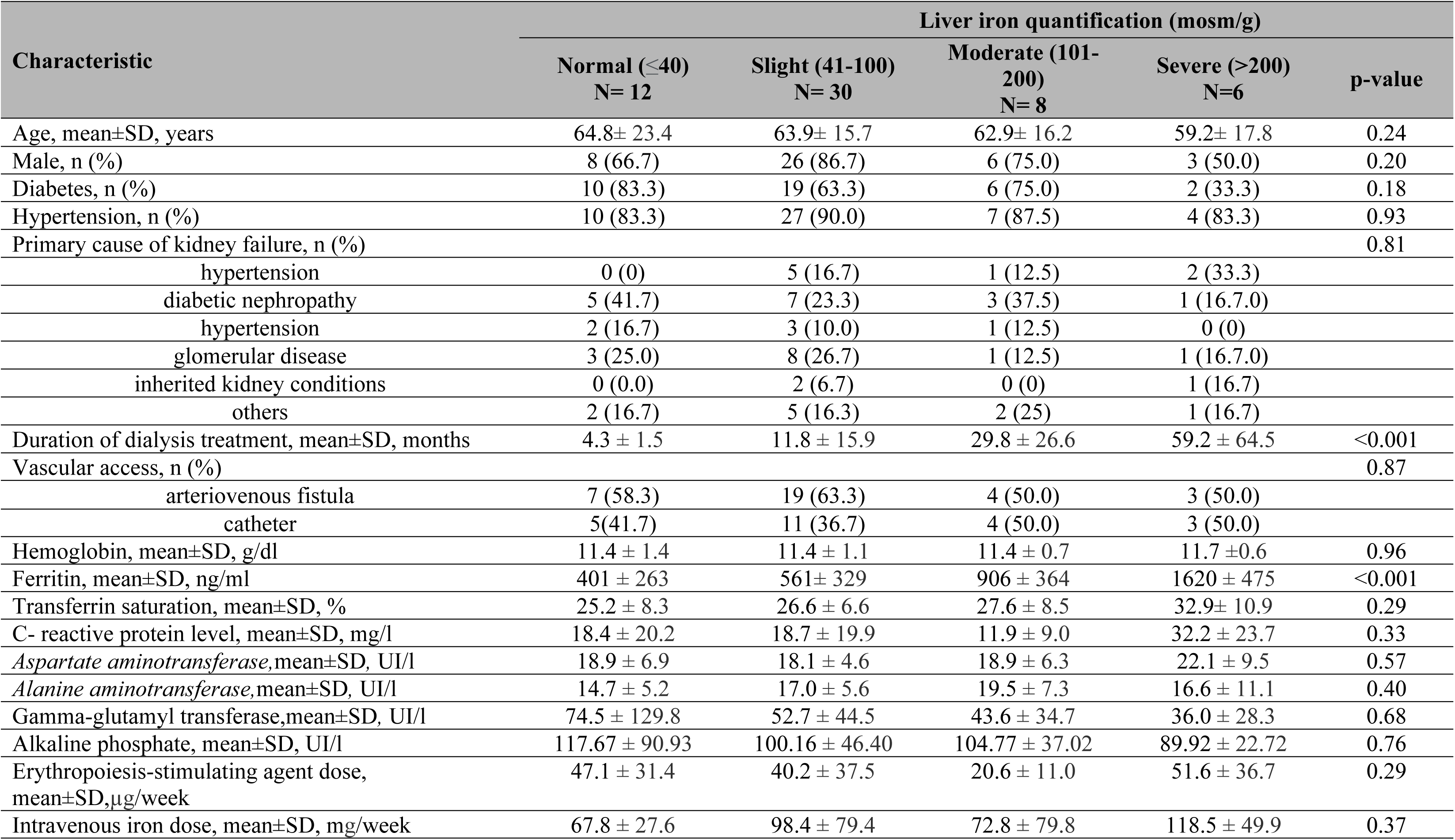
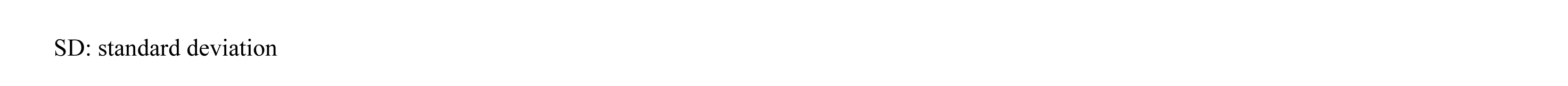
Characteristics of the patients at baseline according to liver iron deposition classified by Rennes score.

| Characteristic | Liver iron quantification (mosm/g) |  |  |  | p-value |
| --- | --- | --- | --- | --- | --- |
| | Normal ($\leq 40$)<br>N= 12 | Slight (41-100)<br>N= 30 | Moderate (101-200)<br>N= 8 | Severe ($>200$)<br>N=6 | |
| Age, mean±SD, years | 64.8± 23.4 | 63.9± 15.7 | 62.9± 16.2 | 59.2± 17.8 | 0.24 |
| Male, n (%) | 8 (66.7) | 26 (86.7) | 6 (75.0) | 3 (50.0) | 0.20 |
| Diabetes, n (%) | 10 (83.3) | 19 (63.3) | 6 (75.0) | 2 (33.3) | 0.18 |
| Hypertension, n (%) | 10 (83.3) | 27 (90.0) | 7 (87.5) | 4 (83.3) | 0.93 |
| Primary cause of kidney failure, n (%) |  |  |  |  | 0.81 |
| hypertension | 0 (0) | 5 (16.7) | 1 (12.5) | 2 (33.3) |  |
| diabetic nephropathy | 5 (41.7) | 7 (23.3) | 3 (37.5) | 1 (16.7.0) |  |
| hypertension | 2 (16.7) | 3 (10.0) | 1 (12.5) | 0 (0) |  |
| glomerular disease | 3 (25.0) | 8 (26.7) | 1 (12.5) | 1 (16.7.0) |  |
| inherited kidney conditions | 0 (0.0) | 2 (6.7) | 0 (0) | 1 (16.7) |  |
| others | 2 (16.7) | 5 (16.3) | 2 (25) | 1 (16.7) |  |
| Duration of dialysis treatment, mean±SD, months | 4.3 ± 1.5 | 11.8 ± 15.9 | 29.8 ± 26.6 | 59.2 ± 64.5 | <0.001 |
| Vascular access, n (%) |  |  |  |  | 0.87 |
| arteriovenous fistula | 7 (58.3) | 19 (63.3) | 4 (50.0) | 3 (50.0) |  |
| catheter | 5(41.7) | 11 (36.7) | 4 (50.0) | 3 (50.0) |  |
| Hemoglobin, mean±SD, g/dl | 11.4 ± 1.4 | 11.4 ± 1.1 | 11.4 ± 0.7 | 11.7 ± 0.6 | 0.96 |
| Ferritin, mean±SD, ng/ml | 401 ± 263 | 561± 329 | 906 ± 364 | 1620 ± 475 | <0.001 |
| Transferrin saturation, mean±SD, % | 25.2 ± 8.3 | 26.6 ± 6.6 | 27.6 ± 8.5 | 32.9± 10.9 | 0.29 |
| C- reactive protein level, mean±SD, mg/l | 18.4 ± 20.2 | 18.7 ± 19.9 | 11.9 ± 9.0 | 32.2 ± 23.7 | 0.33 |
| Aspartate aminotransferase, mean±SD, UI/l | 18.9 ± 6.9 | 18.1 ± 4.6 | 18.9 ± 6.3 | 22.1 ± 9.5 | 0.57 |
| Alanine aminotransferase, mean±SD, UI/l | 14.7 ± 5.2 | 17.0 ± 5.6 | 19.5 ± 7.3 | 16.6 ± 11.1 | 0.40 |
| Gamma-glutamyl transferase, mean±SD, UI/l | 74.5 ± 129.8 | 52.7 ± 44.5 | 43.6 ± 34.7 | 36.0 ± 28.3 | 0.68 |
| Alkaline phosphate, mean±SD, UI/l | 117.67 ± 90.93 | 100.16 ± 46.40 | 104.77 ± 37.02 | 89.92 ± 22.72 | 0.76 |
| Erythropoiesis-stimulating agent dose, mean±SD, μg/week | 47.1 ± 31.4 | 40.2 ± 37.5 | 20.6 ± 11.0 | 51.6 ± 36.7 | 0.29 |
| Intravenous iron dose, mean±SD, mg/week | 67.8 ± 27.6 | 98.4 ± 79.4 | 72.8 ± 79.8 | 118.5 ± 49.9 | 0.37 |

SD: standard deviation

Additionally, we performed a more focused analysis in patients from the two extremes—the first and fourth quartiles—of the hepatic iron deposition distribution quantified by MRI. In this analysis, patients with higher iron deposition upon MRI were on hemodialysis for a longer time; however, there was no difference in the mean weekly dose of intravenous iron therapy and erythropoiesis-stimulating agents in the last 3 months (Table 2). These patients also had higher ferritin levels, with no differences in TSAT, liver toxicity parameters, and C-reactive protein levels between the groups (Table 3).

**Table 2.**
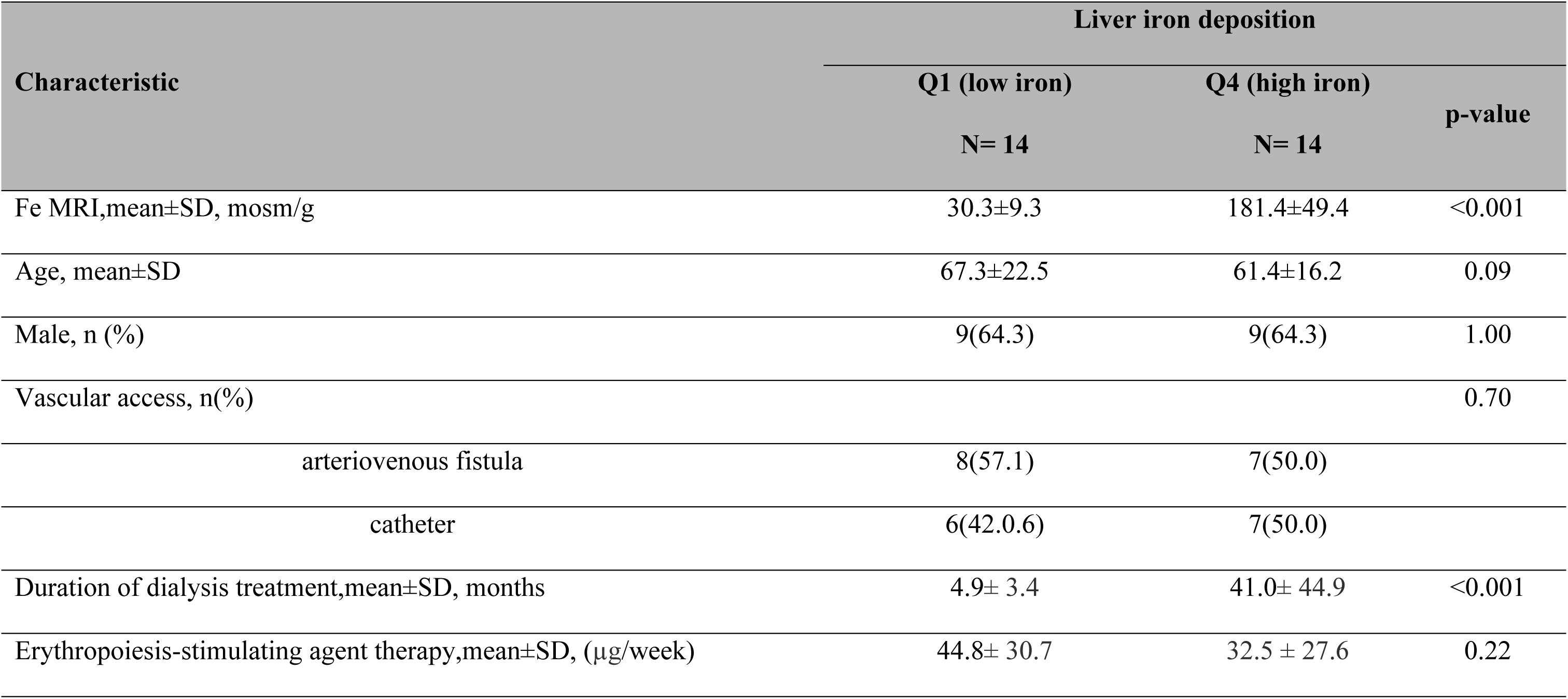
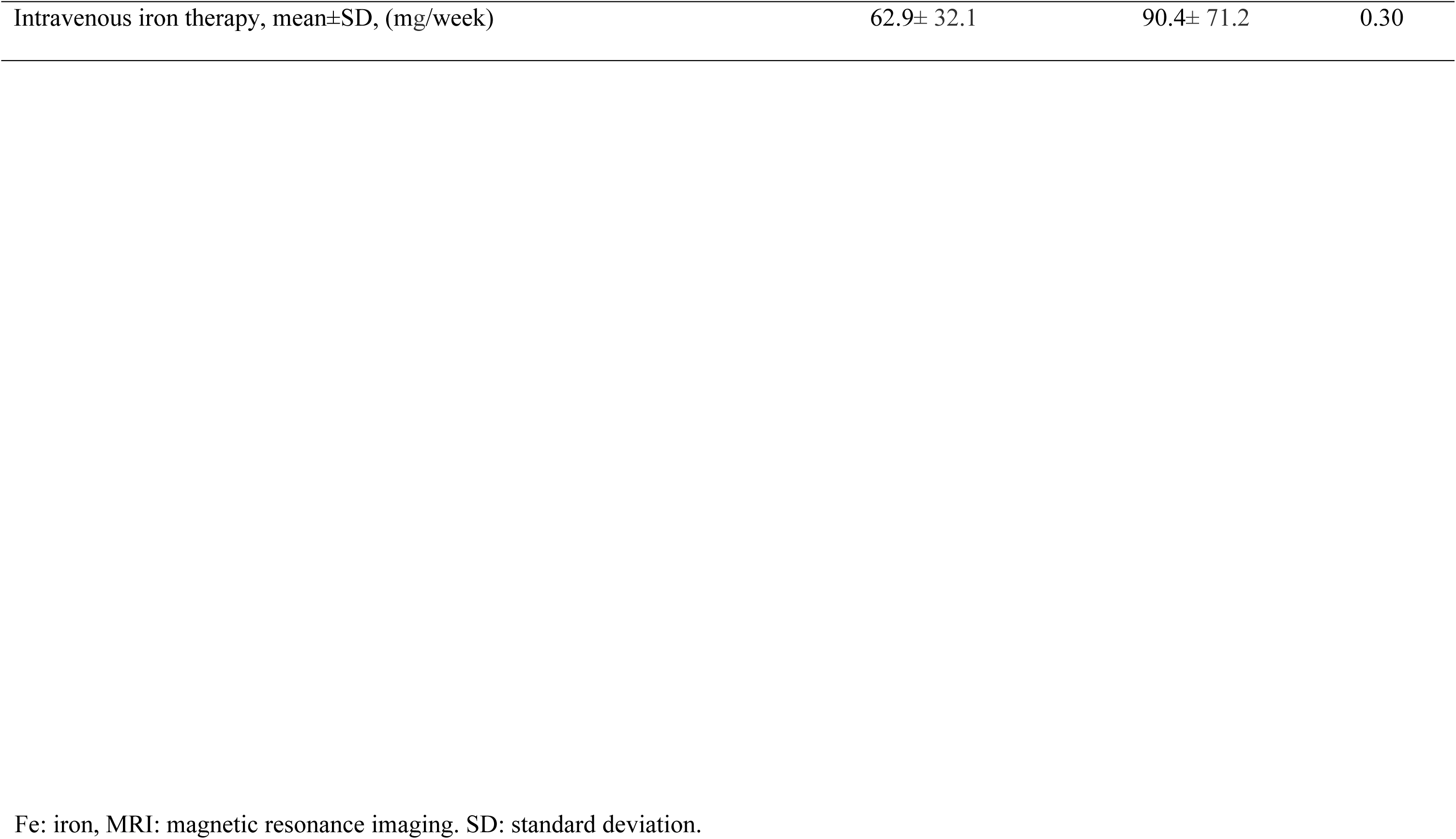
Some baseline characteristics in patients in the first (Q1) and fourth (Q4) quartile, according to iron overload stratified by Rennes score.

| Characteristic | Liver iron deposition |  | p-value |
| --- | --- | --- | --- |
|  | Q1 (low iron)<br>N= 14 | Q4 (high iron)<br>N= 14 |  |
| Fe MRI,mean±SD, mosm/g | 30.3±9.3 | 181.4±49.4 | <0.001 |
| Age, mean±SD | 67.3±22.5 | 61.4±16.2 | 0.09 |
| Male, n (%) | 9(64.3) | 9(64.3) | 1.00 |
| Vascular access, n(%) |  |  | 0.70 |
| arteriovenous fistula | 8(57.1) | 7(50.0) |  |
| catheter | 6(42.0.6) | 7(50.0) |  |
| Duration of dialysis treatment,mean±SD, months | 4.9± 3.4 | 41.0± 44.9 | <0.001 |
| Erythropoiesis-stimulating agent therapy,mean±SD, (µg/week) | 44.8± 30.7 | 32.5 ± 27.6 | 0.22 |
| Intravenous iron therapy, mean±SD, (mg/week) | 62.9± 32.1 | 90.4± 71.2 | 0.30 |

Fe: iron, MRI: magnetic resonance imaging. SD: standard deviation.

**Table 3.**
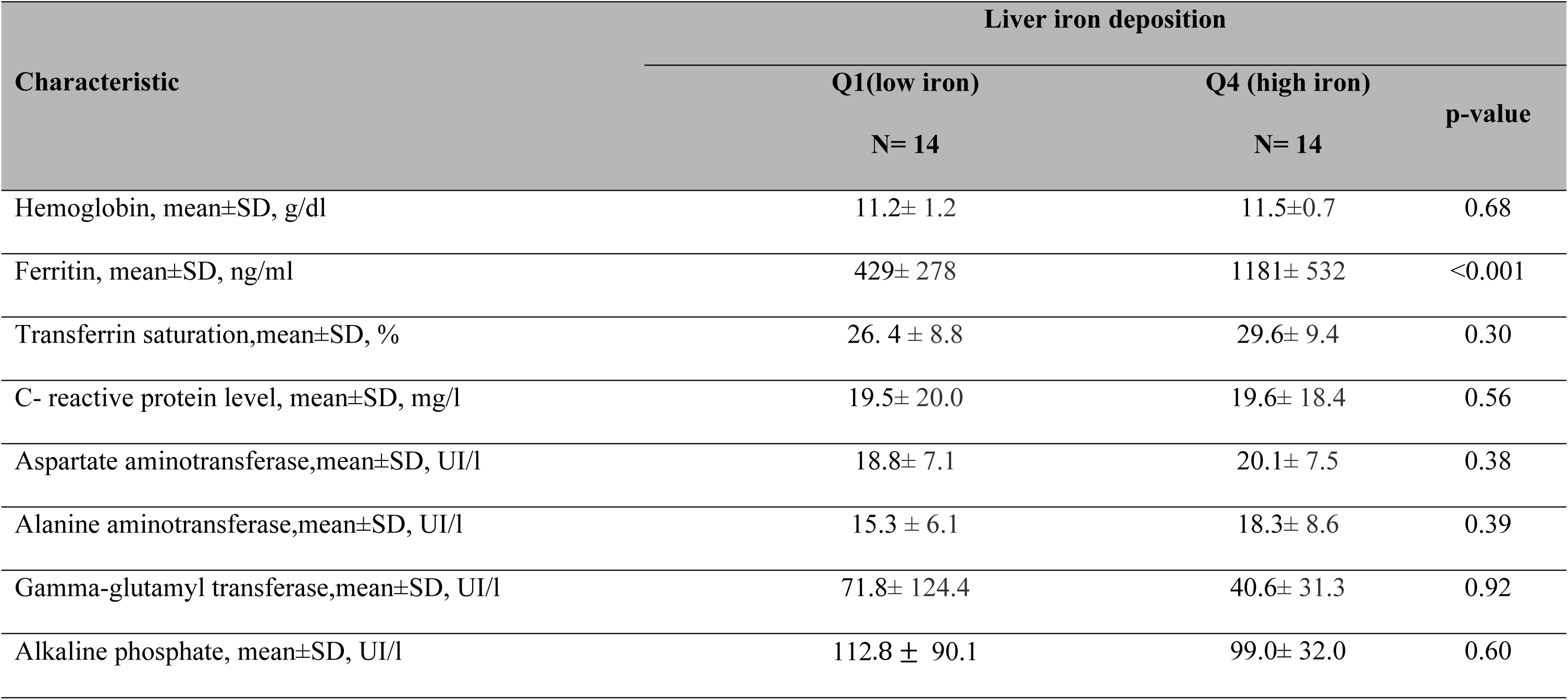
Iron status, systemic inflammation and liver cell function in patients in the first (Q1) and fourth (Q4) quartile, according to iron overload stratified by Rennes score.

| Characteristic | Liver iron deposition |  |  |
| --- | --- | --- | --- |
|  | Q1(low iron) | Q4 (high iron) | p-value |
|  | N= 14 | N= 14 |  |
| Hemoglobin, mean±SD, g/dl | 11.2± 1.2 | 11.5±0.7 | 0.68 |
| Ferritin, mean±SD, ng/ml | 429± 278 | 1181± 532 | <0.001 |
| Transferrin saturation,mean±SD, % | 26. 4 ± 8.8 | 29.6± 9.4 | 0.30 |
| C- reactive protein level, mean±SD, mg/l | 19.5± 20.0 | 19.6± 18.4 | 0.56 |
| Aspartate aminotransferase,mean±SD, UI/l | 18.8± 7.1 | 20.1± 7.5 | 0.38 |
| Alanine aminotransferase,mean±SD, UI/l | 15.3 ± 6.1 | 18.3± 8.6 | 0.39 |
| Gamma-glutamyl transferase,mean±SD, UI/l | 71.8± 124.4 | 40.6± 31.3 | 0.92 |
| Alkaline phosphate, mean±SD, UI/l | 112.8 ± 90.1 | 99.0± 32.0 | 0.60 |

Moreover, no differences were found between these two groups in PBMCs iron content (Table 4) or in the plasma content of HNE protein adducts (Figure1).Regarding the inflammatory parameters assessed in PBMCs, no differences were found for MCP-1 and TNF-α; however, lower IL-6 levels were detected in patients with higher hepatic iron deposition(Figure 1).

**Figure 1.**
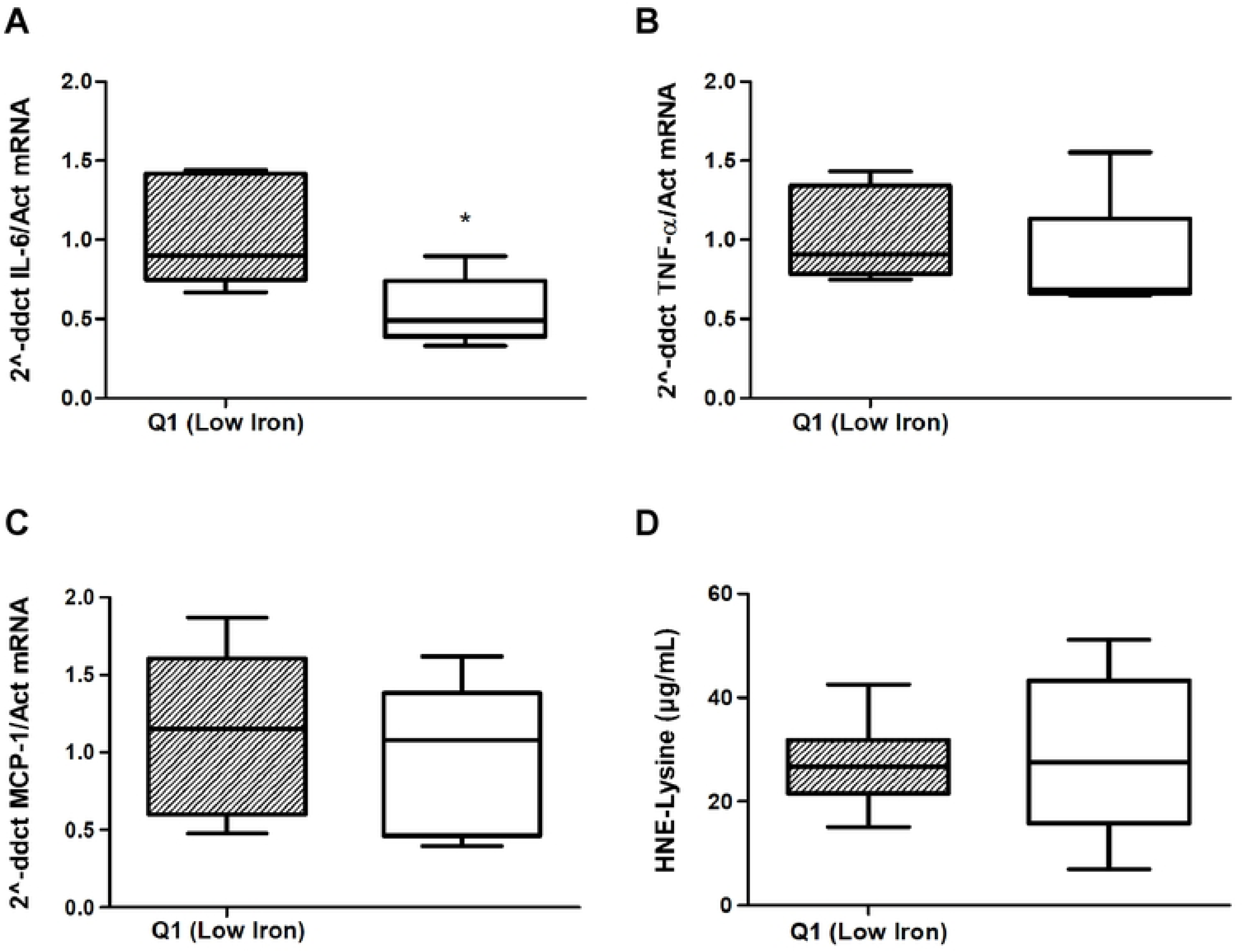
Proinflammatory cytokine mRNA content in peripheral blood mononuclear cells and (hydroxynonenal) HNE protein adducts in plasma in patients in the first (Q1) and fourth (Q4) quartile, according to iron overload stratified by Rennes score. IL-6: Interleukin 6, TNF-α: tumor necrosis factor alpha, MCP-1: monocyte chemoattractant protein-1, HNE-Lysine: 4-hydroxynonenal- lysine.*p<0.05. Regarding the inflammatory parameters assessed in PBMCs, no differences were found for TNF-α and MCP-1 (Figure 1B and 1C), but lower IL-6 levels were detected in patients with higher hepatic iron deposition (Figure 1A). No differences were found in the adducted plasma content of the HNE protein (Figure 1 D).

**Table 4.** Quantification of iron content in peripheral blood mononuclear cells in patients in the first (Q1) and fourth (Q4) quartile, according to iron overload stratified by Rennes score.

| Characteristic | Liver iron deposition |  |  |
| --- | --- | --- | --- |
|  | Q1 (low iron) | Q4 (high iron) | p-value |
|  | N= 14 | N= 14 |  |
| Total iron, median (interquartile range)- (nmol/mg protein) | 0.48 (0.77) | 1.03 (0.87) | 0.24 |
| Ferrous iron($\text{Fe}^{2+}$), median (interquartile range)- (nmolFe/mg protein) | 0.30(0.45) | 0.47(0.45) | 0.18 |
| Ferric iron ($\text{Fe}^{3+}$), median (interquartile range)- (nmolFe/mg protein) | 0.21 (0.74) | 0.38(0.50) | 0.14 |

## Discussion

Intravenous iron therapy is useful for improving anemia in hemodialysis patients. Increasing the total amount of iron administered can reduce the requirement for erythropoiesis-stimulating agents ^(13)^. However, there are concerns regarding the potential toxicity of iron overload. The KDIGO2012 Anemia guideline recommends the administration of intravenous iron with TSAT levels ≤30% and ferritin ≤500µ/l^(2)^. However, these recommendations are mainly based on observational studies, and the safe upper limits for TSAT and ferritin remain unknown^(4)^.

Considering the available information, we have some doubts regarding the relevance of ferritin concentrations in prescribing iron therapy. Ferritin is not a good parameter to assess iron availability for erythropoiesis, and hemoglobin and TSAT levels alone may be more appropriate for deciding how to administer intravenous iron. Moreover, we question the evidence that the iron sequestered in the liver or circulating immune cells of hemodialysis patients could induce significant tissue damage or facilitate infection by pathogenic bacteria, since it is not free in blood circulation. Iron is a transition metal that is highly active in oxidation-reduction reactions, and it must be bound to a ligand to avoid damage to cells and tissues. These ligands mainly include transferrin, heme, and ferritin. Iron not bound to these ligands, called non-transferrin bound iron (NTBI), appears to be responsible for damage both on cell surfaces and intracellularly ^(18–19)^ and may be a risk factor for gram-negative and other siderophilic bacteria ^(8–9)^.

In this study, we evaluated the potential toxicity of iron deposition in prevalent hemodialysis patients receiving intravenous iron therapy, as well as its relationship with the parameters used to assess iron status (ferritin and TSAT). We found that patients with higher hepatic iron overload were those with a longer time on hemodialysis; however, they did not differ in the current mean weekly iron and erythropoiesis-stimulating agent therapy, suggesting that the differences found could be better explained by a higher cumulative iron dose over time. Similarly, patients with higher iron deposition had higher ferritin levels in the absence of significant differences in TSAT, supporting the opinion that ferritin is a good marker of hepatic iron deposition. However, regarding iron deposition in other organs, MRI studies have shown that iron deposition in organs other than the liver, such as the heart or pancreas, has a different pattern and is not correlated with serum ferritin levels ^(20–23)^. Our results support these findings, as no differences were observed in the iron content of PBMCs between the two experimental groups tested. Therefore, our results indicate that only TSAT values agree with the KDIGO recommendations, and this is due to doubts regarding the clinical value of ferritin as a good marker for anemia treatment in hemodialysis patients.

Regarding organ and cell toxicity, we were unable to detect significant changes in the parameters tested as a function of hepatic iron accumulation or ferritin levels. Additionally, the circulating levels of hepatic enzymes, a generally accepted marker of liver toxicity, the concentration of circulating oxidized proteins, and the pattern of synthesis of MCP-1 and TNF-α by PBMCs, did not differ among the patients studied. This lack of toxicity may be related to the mechanisms involved in cellular iron homeostasis. For example, hepcidin plays an important role in anemia in patients with chronic kidney disease (CKD) by inducing functional iron deficiency. In CKD, different circumstances converge to increase hepcidin production in the liver, such as a proinflammatory state, decrease in erythropoietic activity, and decrease in hepcidinclearance^4^.Hepcidin blocks the activity of ferroportin, a transmembrane protein that allows iron release for use within cells. Ferroportin is mainly found in intestinal epithelial cells and macrophages and inhibits the efflux of iron from these cells into the plasma, promoting its sequestration in the liver ^(24)^.In diseases with iron overload, such as thalassemia, hemochromatosis, or severe liver disease, hepcidin is decreased. In these cases, excess circulating iron may increase in the form of NTBI ^(18)^. The toxicity in these pathologies appears to be caused by NTBI, which can lead to lipid peroxidation, endothelial dysfunction, and organ and tissue damage ^(25–26)^.NTBI increases significantly when TSAT values rise above 70%; however, it does not correlate with serum ferritin levels ^(19)^.Therefore, TSAT may be a better marker of available iron for erythropoiesis than ferritin.

As previously stated, MCP-1 and TNF-α levels were comparable in the PBMCs of patients with high and low hepatic iron deposition and ferritin levels; however, the IL-6 levels in PBMCs decreased in patients with increased hepatic iron deposition. In fact, we noted that individuals with a higher deposition had lower IL-6 levels. The interpretation of this finding is unclear, as IL-6 is a pro-inflammatory cytokine, and its decrease could have beneficial effects. However, it plays an important role in the immune response; therefore, its deficiency could potentially affect this.

Our study had some limitations. First, this was a cross-sectional study; therefore, causality could not be demonstrated. In addition, the number of patients included in the study was limited. Although a previous randomized clinical trial has shown that intravenous iron therapy in hemodialysis patients with a high ferritin limit of 700 μg /l ^(13)^. is safe, further studies are necessary to confirm this safety at higher ferritin levels.

In summary, in hemodialysis patients treated with intravenous iron and ferritin levels higher than those recommended by clinical practice guidelines, our results do not support the appearance of increased liver toxicity or proinflammatory and oxidative damage. Therefore, we believe that hemodialysis patients are likely to benefit from receiving higher iron doses when it is considered useful for treating anemia without an increased risk of toxicity, and that the available markers have limitations in determining the risk of toxicity. In the future, we may have new tools to assess iron toxicity, such as the NTBI, which may facilitate treatment decisions^(19)^.Additionally, new treatments involving hepcidin to improve iron bioavailability, such as inhibitors of hypoxia-inducible factor and prolyl hydroxylase or monoclonal antibodies targeting the hepcidin-ferroportin pathway, are currently being developed ^(27–28)^. Until these new drugs can be widely used, we believe that the benefit of more proactive doses of iron outweighs the risk of toxicity and should be considered when treating anemia in hemodialysis patients.

## Data Availability

All relevant data are within the manuscript and n Supporting Information files.

## Acknowledgements

The Ethics Committee of the Hospital Universitario Príncipe de Asturias, considered issuing a favorable opinion for the study to be carried out. Conflict of Interest Statement: none declared.

